# Integrating DHS/MIS Biomarkers with 34 Years of CHIRPS-NDVI Climate Data for Malaria Risk Prediction in Nigeria: A Machine Learning and Spatial Mapping Approach

**DOI:** 10.64898/2025.12.14.25342244

**Authors:** Daniel Onimisi

## Abstract

**Background:** The estimates of national disease risk are considerably limited by the time of conducted surveys and the geographical inadequacies in surveillance, notwithstanding malaria’s continued prominence in morbidity and mortality in Nigeria. There is limited research employing machine learning to integrate long term environmental trends with DHS/MIS biomarker data on a national scale, despite the established influence of climate, rainfall patterns, vegetation, and population on transmission dynamics.

Nigeria has one of the highest rates of malaria cases in the world, and the disease is still a major public health problem there. Climate variability greatly affects malaria transmission; however, comprehensive national studies that combine DHS/MIS biomarker data with climate and vegetation dynamics to accurately forecast spatial malaria risk at a high resolution are lacking.

This research presents a hybrid framework that integrates machine learning with geospatial analysis, utilizing DHS/MIS survey biomarkers from 2010, 2015, and 2021, alongside CHIRPS rainfall data, MODIS-NDVI vegetation indices, and climate-trend features to forecast malaria risk at the LGA level throughout Nigeria.

**Methods:** We linked a thorough MIS biomarker dataset (n = 139,407) with LGA boundaries and added climatic variables such rainfall, NDVI, temperature, population, long-term climate averages, anomalies, and indicators that were one and three years behind. We used data from three MIS years to create a random forest classifier and tested its capacity to work across multiple survey periods by using cross-year validation. We used feature significance and SHAP explainability techniques to look at how the model worked.

Malaria risk maps for all of Nigeria have been made to help people make decisions. They have a resolution of 1 to 5 kilometers and are arranged by LGA and state administrative boundaries. At the LGA level, malaria risk maps were made that showed risk categories from 0 to 7, as well as surfaces based on chance.

**Results:** Our hybrid model predicted RDT-based infection status with cross-year validation accuracies of 62.2% (2010), 36.8% (2015), and 40.2% (2021), reflecting temporal shifts in climate, vector, and intervention dynamics. SHAP analyses identified ITN coverage, 3-year climate lag rainfall/NDVI means, temperature, population density, and rainfall variability as dominant predictors, revealing strong linkages between vegetation greenness, rainfall patterns, and malaria prevalence.

LGA-level spatial mapping generated national probability surfaces and risk stratifications (0 to 7%), confirming the existence of persistent hotspots in the northern savannah and river-basin regions, as well as climate-related elevated risks in North-Central and South South LGAs, due to anomalous rainfall and accelerated vegetation regeneration. These patterns align with observed MIS prevalence trends across all three survey years, illustrating the robustness of the model and the advantage of integrating 34 years of environmental time series. Our hybrid model predicted RDT-based infection status with cross-year validation accuracies of 62.2% (2010), 36.8% (2015), and 40.2% (2021), indicating temporal fluctuations in climate, vector, and intervention dynamics. SHAP analysis found that ITN coverage, a three-year climatic lag of rainfall/NDVI averages, temperature, population density, and rainfall variability are key predictors, demonstrating substantial relationships between vegetation greenness, rainfall patterns, and malaria prevalence.

**Conclusion:** This study illustrates that the integration of Demographic and Health Survey/Malaria Indicator Survey (DHS/MIS) biomarkers with 34 years of Climate Hazards Group InfraRed Precipitation with Station data (CHIRPS) rainfall and Moderate Resolution Imaging Spectroradiometer Normalized Difference Vegetation Index (MODIS NDVI) vegetation data establishes a robust, scalable machine-learning framework for forecasting malaria risk in Nigeria. The model precisely delineates climate related epidemiological and environmental parameters, generates risk surfaces at the state and local government area levels consistent with MIS distributions, and finds hotspots of climatic anomalies that exacerbate transmission.

This replicable digital epidemiology framework enables operational early warning systems, subnational targeting, resource distribution, public health monitoring, and climate-resilient control strategies. Future study will expand forecasts to include 2024 to 2035 by employing climate projections and incorporating DHS data from 1990 to 2024 to enhance temporal validity.

Data sources: DHS/MIS biomarker data; CHIRPS precipitation; NDVI derived from satellite remote sensing products accessed via Google Earth Engine.

## 1. Introduction

The use of insecticide-treated nets (ITNs), seasonal malaria chemoprevention (SMC), and improved diagnostics has not alleviated the widespread problem of malaria in Nigeria, which is responsible for about 27% of the world’s cases and 31% of malaria deaths (WHO, 2023). Transmission is affected by a complex web of interrelationships between population trends, intervention coverage, climate variability (including temperature, precipitation, and vegetation), and ecological zones (Bhatt et al., 2015; Ryan et al., 2020).

Temperature, precipitation, and vegetation indices have historically been used as proxy indicators to forecast mosquito habitat suitability using climate-centric statistical methods (Midega et al., 2018; Shapiro et al., 2017).

Despite the fact that CHIRPS rainfall and MODIS-NDVI datasets enable detailed environmental monitoring (Funk et al., 2015) and that DHS/MIS surveys generate reliable biomarker-validated parasitemia, ITN utilization, and geocoded clusters (NPC & ICF, 2019; NMIS, 2021). However, traditional models seldom incorporate these supplementary data sources. Previous machine learning approaches have their drawbacks, such as climate-exclusive predictions, single-year frameworks, and a lack of LGA-level outputs that are necessary for programmatic interventions (Adewoyin et al., 2022; Midega et al., 2018). This means that even when demographic and behavioral characteristics are measured in surveys that are typical of the population as a whole, climate-only prediction models don’t always account for fluctuations in these variables.

The Demographic and Health Surveys/Malaria Indicator Surveys (DHS/MIS) provide information on household characteristics, geocoded sampling clusters, ITN utilization, and biomarker-verified parasitemia. According to the National Malaria Elimination Programme [NMEP], 2021, these surveys provide the most reliable ground-truth data for estimating Nigeria’s malaria burden over several years. Despite the potential to improve spatial accuracy, model interpretability, and policy applicability, few studies have successfully integrated DHS/MIS biomarker data with long-term climate–vegetation time series.

With its improved capacity to handle nonlinear ecological relationships, large feature sets, and spatial heterogeneity, machine learning (ML) has become a more and more useful approach for malaria prediction (Baratloo et al., 2022; Masinde, 2020). Earlier machine learning research, both globally and in Nigeria, has shown encouraging results using climate or sociodemographic factors (Adewoyin et al., 2022; Gething et al., 2011); however, it often faces one or more constraints:

1. The analysis lacks biomarker-validated prevalence data and only uses climatic records.
2. Single-year or single-survey designs restrict temporal generalizability.
3. ito life cycles, are not sufficiently incorporated into the analysis.
4. Geographical outputs, which are essential for making wise programming choices, are lacking at the LGA level.

To address these gaps, we present a hybrid modelling framework that integrates DHS/MIS biomarker data (2010, 2015, 2021) with 34 year rainfall–NDVI climate features, including multi-year means, anomalies, and lag structures derived from long-term CHIRPS and satellite vegetation datasets. Using a machine learning approach, we generate malaria risk maps at the LGA level and evaluate model performance using cross-year temporal validation.

The study aims to:

1. Construct a harmonized dataset combining biomarker-confirmed malaria results with climate–vegetation variables across three national MIS years.
2. Train and validate a machine learning model for malaria risk classification using both socio-demographic and environmental predictors.
3. Generate high-resolution spatial risk maps to support malaria program decision-making at LGA and state levels.
4. Interpret model behaviour using feature importance and SHAP explainability techniques.

This study pioneers the first multi-year machine learning model in Nigeria directly integrating DHS/MIS parasitemia biomarkers (2010, 2015, 2021) with 34 years (1990–2024) of CHIRPS rainfall, MODIS-NDVI, and environmental covariates processed via Google Earth Engine. The random forest classifier enhanced with lagged/anomaly climate features, as well as SHAP interpretability, generates LGA-level risk maps through cross-year validation, demonstrating feasibility for subnational surveillance in non-survey years while enhancing explainability and predictive robustness.

Spatial analyses demonstrate significant heterogeneity, with enduring high-burden clusters in the northern Sahelian belt and Middle Belt, contrasted by lower-risk southern coastal zones. Climate anomaly mapping reveals a significant correlation between elevated malaria risk and regions characterized by high humidity and temporal variations in vegetation. This indicates that climate change facilitates the transmission of the disease.

This biomarker-grounded, climate-aware digital pipeline identifies key environmental drivers and hotspots, equipping Nigeria’s National Malaria Elimination Programme with a scalable platform for targeted interventions, rapid response, early warning systems, and climate-resilient public health planning.

## 2. METHODS

### 2.1 Study Design and Overview

This research effort entailed the creation and validation of a predictive machine-learning framework aimed at evaluating malaria risk across Nigeria. The objective was to delineate malaria risk patterns throughout Nigeria and produce spatially continuous risk maps at the Local Government Area (LGA) level.

1. The first step involved extracting and harmonizing malaria biomarker variables (RDT results) from the MIS datasets.
2. The next step involved merging these with cluster-level GPS and climate/vegetation covariates.
3. The goal was to train machine learning (ML) models to predict cluster-level malaria risk and create national and local government area (LGA) risk maps.
4. The models were then validated using cross-year holdouts from the Malaria Indicator Surveys (MIS) conducted in 2010, 2015, and 2021.

### 2.2 Data Sources

Malaria Indicator Survey (MIS) datasets for Nigeria were obtained for the survey years 2010, 2015 and 2021 from the DHS program repository. This methodology amalgamated household biomarker data from Malaria Indicator Surveys (MIS/DHS) with longitudinal climate and vegetation time-series data (rainfall and NDVI) gathered from 1990 to 2024. Individual and cluster level variables included rapid diagnostic test(RDT) outcomes, insecticide-treated net (ITN) coverage, prior malaria prevalence, population estimates, and georeferenced cluster coordinates displaced according to the DHS privacy protocols.

Thus, RDT results served as the primary outcome variable categorized into ordinal malaria risk classes (0-7) representing increasing malaria burden. Survey clusters were spatially linked to the administrative boundaries using geographic coordinates.

The analytical workflow also included harmonization, climate feature engineering, geospatial integration, model training, cross-year temporal validation, SHAP model interpretation, and visualization of national risk maps.

#### 2.2.1 DHS/MIS Biomarker Data (2010, 2015, 2021)

We utilized georeferenced household cluster datasets from the DHS/MIS rounds corresponding to years with nationally representative biomarker surveys. Datasets were extracted in Stata format (.DTA) and subsequently processed into standardized CSV files.

● MIS/DHS survey data (biomarkers): MIS datasets for Nigeria encompassing MIS years 2010, 2015 and 2021 were downloaded from the DHS Program and stored under data/mis_datasets/. RDT and related variables were identified programmatically from .DTA and .DO files and extracted into outputs/MIS_Malaria_Extracted.csv and R-extracted MIS_Malaria_Extracted_R.csv. The minimal RDT dataset used for modelling is outputs/MIS_Malaria_RDT_Minimal.csv.gz and the cleaned/tested dataset is outputs/MIS_Malaria_RDT_Clean.csv.gz. (Counts: minimal RDT n ≈ 19,629; combined dataset after merges ≈ 139,407 rows; training rows ≈ 137,848).
● GPS / cluster shapefiles: DHS cluster GPS shapefiles for MIS years were read from data/mis_datasets/*/NGGE*FL/*.shp and joined to the extracted RDT data via cluster ID (DHSCLUST / hv001). GPS displacement/readme files were examined and noted for limitations.
● Climate and vegetation time-series: CHIRPS rainfall and NDVI (from MODIS or processed NDVI layers) covering 1990–2024 were preprocessed and merged to cluster-level timeseries. The merged national rainfall file is outputs/Nigeria_CHIRPS_Rainfall_1990_2024.csv and NDVI products are in outputs/Nigeria_DHS_NDVI_1990_2024_FULL.csv and related files.

#### 2.2.2 Preprocessing and variable extraction

● **RDT extraction from survey DTA:** We scanned .DO/.DTA files to find malaria-related variable names (e.g., ml*, hml*, rdt*, smalar). For each survey file, the candidate RDT columns were validated and the valid columns concatenated into long format (outputs/MIS_Malaria_RDT_Long.csv/mis_rdt_final.csv). A minimal standardized RDT column rdt_result (0/1) was derived. Child index / record IDs were retained.
● **Metadata extraction:** The script pulled cluster/region/weight variables (hv001 / v001 / DHSCLUST, region fields, survey weights where available) into a metadata table and merged it with the RDT minimal data. Final merged metadata combined rows ≈ 238,762 (intermediate) and final modeling table ≈ 139,407 rows after deduplication and joins.
● **GPS join:** Cluster shapefiles were read (using geopandas), converted to latitude/longitude columns (LATNUM / LONGNUM) and merged on year + cluster id; results saved as outputs/mis_rdt_with_gps.csv. Spatial joins handled column-name conflicts (dropped index_right if present). Displacement README files were consulted and displacement constraints noted.
● **Climate/NDVI merging and feature engineering:** For each cluster and year we computed climate/NDVI features aggregated to cluster scale from the 1990–2024 timeseries. Key derived variables used by the model include:

○ Rainfall (annual or seasonal aggregate aligned to survey)

○ NDVI (cluster mean for the same period)

○ Temperature (cluster mean)

○ Population (cluster population covariate from DHS/GIS)

○ ITN (insecticide treated net coverage proxy from survey covariates b5/b6 or ITN_Coverage fields)

○ Prev (previous prevalence where available)

○ **Climate time features** derived from the 34-year time series: Climate_mean (long-term climatology), climate_lag1 (year lag), climate_lag3_mean (3-year rolling mean), Climate_baseline (multi-year baseline), and climate_anomaly (current minus baseline). The feature engineering script saves outputs/mis_climate_features.csv. (Note: rolling/lag were computed by groupby([’State’,’LGA’]).transform(rolling(…)) to avoid index misalignment.)

Primary outcome was binary RDT positivity (rdt_result). For map visualizations we converted model probabilities into ordinal risk classes (0–7) then labelled classes (Very Low → Very High). Key extracted variables included:

● RDT malaria positivity (binary outcome)
● DHS geocoordinates (lat/long)
● Household and individual variables:

○ Insecticide-treated net use (ITN)

○ Fever prevalence proxy (Prev)

○ Population estimates (Population)

#### 2.2.3 Climate & Vegetation Time Series (CHIRPS & MODIS NDVI) Data Acquisition

Long-term climate and vegetation variables were retrieved using Google Earth Engine(GEE), which enabled scalable procesing of multi-decadal satellite imagery. Vegetation dynamics were characterizedd using the Normalized Difference Vegetation Index (NDVI) derived fromMODIS (Moderate-resolution Imaging Spectroradiometer) which refers to NASA’s advanced Earth-observing sensors on the Terra (1999) and Aqua (2002) satellites and Landsat while precipitation was obtained from the CLimate Hazards Group InfraRed Precipitation with Stations (CHIRPS) dataset.

For each MIS cluster location, rainfall and NDVI values were extracted from long-term satellite archives:

● CHIRPS rainfall (0.05° resolution)
● MODIS NDVI (16-day composites)

Monthly NDVI and rainfall data were extracted for the period **1990–2024**, spatially aggregated over Nigeria, and temporally summarized to generate annual climatologies. These long-term datasets were used to derive contextual climate features that complemented the MIS survey years.

We generated annual climate summaries for each MIS year (2010, 2015, 2021):

● Annual Rainfall (mm)
● Annual mean NDVI
● Annual mean Temperature (from climate reanalysis)

### 2.3 Climate Feature Engineering

To capture interannual variability and climate-driven malaria suitability, we created five engineered features:

1. Climate_mean Average rainfall/NDVI composite for the MIS year.
2. climate_lag1 One-year climate lag to incorporate delayed ecological effects of rainfall on vector breeding cycles.
3. climate_lag3_mean The three-year rolling mean represents a smoothed trend in climate suitability.
4. Climate_baseline Long-term mean (climatological baseline) from historical climate for each location.
5. climate_anomaly

Difference between MIS-year climate and baseline — proxy for climate shocks.

All features were validated to be continuous, non-collinear, and predictive in prior literature. They also captured both contemporaneous and delayed climatic influences on malaria transmission dynamics.

### 2.4 Spatial Data Processing and Integration

All datasets were projected to a common coordinate reference system (EPSG:4326). Spatial joins were validated to prevent duplication or boundary conflicts.

The final analytical dataset consisted of 137,848 georeferenced observations with demographic, epidemiological, and climate-vegetation attributes across Nigeria. This section describes the spatial data processing workflow employed to ensure coordinate accuracy, administrative boundary alignment, and compatibility for subsequent geospatial analysis and visualization.

Unlike recent geospatial ML studies that rely exclusively on environmental predictors (e.g., [IJGI, 2024]), this study integrates DHS/MIS biomarker-derived malaria outcomes with long-term climate–vegetation dynamics, enabling epidemiologically grounded risk prediction and cross-survey validation.

#### 2.4.1 Spatial Cleaning and Coordinate Standardization

All cluster GPS coordinates underwent systematic processing to address common geospatial data quality issues. First, coordinates were standardized to a consistent coordinate reference system using the WGS84 datum (EPSG:4326), ensuring compatibility across all spatial datasets.

Second, erroneous entries including zero coordinates and duplicate location records were identified and removed from the dataset. Third, we accounted for the inherent spatial displacement introduced by DHS GPS masking protocols, which intentionally jitter cluster coordinates by up to 2 kilometers in urban areas and up to 5 kilometers (with 1% displaced up to 10 kilometers) in rural areas to protect respondent confidentiality. While these offsets could not be reversed, awareness of this uncertainty informed subsequent spatial analysis decisions, particularly regarding the precision of cluster-to-administrative unit assignments.

#### 2.4.2 Local Government Area Linking and Administrative Boundary Integration

To enable aggregation and visualization of malaria predictions at administratively relevant scales, we linked each MIS cluster to its corresponding Local Government Area using GADM version 4.1 administrative boundaries. Nigeria comprises 774 LGAs, which serve as the third-level administrative divisions below states and represent meaningful units for public health planning and resource allocation. The spatial join was executed using the geopandas.sjoin function, which assigned each cluster point to the LGA polygon within which it fell. During this process, we identified and programmatically resolved conflicts arising from "index_right" artifacts—spurious duplicate assignments that can occur when points fall near polygon boundaries or when multiple geometries overlap. These conflicts were systematically removed to ensure each cluster mapped to exactly one LGA. This linkage enabled subsequent aggregation of individual-level predictions to LGA-level prevalence estimates and facilitated the generation of choropleth maps displaying spatial patterns of malaria risk across Nigeria’s administrative landscape.

#### 2.5.1 Outcome Variable Definition and Risk Stratification

The primary outcome variable was laboratory-confirmed malaria infection status, ascertained through rapid diagnostic test (RDT) results coded as a binary indicator (0 = negative, 1 = positive). To facilitate epidemiologically interpretable risk stratification and enable targeted intervention planning, individual-level RDT positivity was aggregated and transformed into categorical malaria risk classes ranging from 0 to 7. This ordinal classification scheme was constructed by calculating cluster-level or area-level prevalence estimates and applying epidemiologically informed thresholds that partition the prevalence continuum into discrete transmission intensity strata.

The numeric risk classes (0–7) were subsequently mapped to descriptive epidemiological labels aligned with World Health Organization malaria transmission classification frameworks and established endemicity terminology. The eight-level stratification scheme encompassed: Very Low (class 0), Low (class 1), Low-Medium (class 2), Moderate (class 3), High (class 4), Very High (class 5), Severe (class 6), and Extreme (class 7) transmission categories. This granular classification preserves meaningful epidemiological distinctions across the full spectrum of malaria endemicity observed in Nigeria, ranging from areas approaching pre-elimination status to hyperendemic zones with sustained year-round transmission. The categorical outcome structure enables both multi-class machine learning classification and clear communication of spatially heterogeneous malaria burden to public health stakeholders responsible for resource allocation and intervention targeting decisions.

#### 2.5.2 Feature Set

The final predictor variable set comprised eleven features spanning environmental, climatic, demographic, and intervention domains, selected based on established epidemiological theory, bivariate associations with malaria outcomes, and iterative feature importance assessments during preliminary model development. This multidimensional feature space captures the principal ecological and anthropogenic determinants of malaria transmission dynamics in Nigeria.

##### Environmental and Climatic Variables

Rainfall (mm) and temperature (°C) were included as direct measures of hydroclimatic conditions governing Anopheles mosquito larval habitat availability, adult mosquito survival, and parasite developmental rates within the vector. The Normalized Difference Vegetation Index (NDVI) served as a proxy for vegetation density and land cover characteristics that influence mosquito breeding site availability and microclimate suitability for vector populations. To characterize temporal dynamics in climate-malaria associations, several derived climate variables were constructed: climate_mean represents the long-term average climatic conditions at each location; climate_lag1 and climate_lag3_mean capture lagged climatic exposures at one-month and three-month intervals respectively, reflecting the biologically plausible delay between environmental conditions and subsequent malaria case manifestation; climate_baseline establishes the historical climatological norm for each location; and climate_anomaly quantifies deviation from expected conditions, enabling detection of abnormal climatic events that may trigger epidemic transmission.

##### Intervention and Demographic Variables

Insecticide-treated net (ITN) coverage represented the primary vector control intervention, measured as the household-level proportion of the population sleeping under ITNs during the night preceding the survey. Fever prevalence (Prev) served as a proxy indicator for recent febrile illness burden within the household, capturing both malaria-attributable fever and broader health system stress that may correlate with transmission intensity. Population density (persons per square kilometer) characterized human settlement patterns and the concentration of susceptible hosts, which influences force of infection through effects on vector-human contact rates and healthcare access.

This integrated predictor set aligns with the feature importance rankings derived from SHAP analysis and represents the complete set of model inputs employed during Random Forest classifier training, ensuring consistency between feature selection rationale, model implementation, and interpretability assessments reported in subsequent sections.

Final predictor set included:

● Rainfall
● NDVI
● Temperature
● ITN coverage
● Prev (fever proxy)
● Population density
● Climate_mean
● climate_lag1
● climate_lag3_mean
● Climate_baseline
● climate_anomaly

These matched the feature importance results and model inputs used during training.

#### 2.5.3 Algorithm Selection and Model Development

##### Comparative Algorithm Evaluation

The selection of an appropriate machine learning algorithm was informed by systematic comparison of multiple candidate modeling frameworks, each offering distinct advantages for epidemiological prediction tasks. Four algorithms were initially evaluated for their capacity to capture complex, non-linear relationships between predictor variables and categorical malaria risk outcomes:

1. **Logistic Regression**: A generalized linear model providing interpretable coefficient estimates and probabilistic predictions, but limited in its capacity to capture non-linear feature interactions without explicit polynomial or interaction term specification.
2. **Random Forest**: An ensemble method constructing multiple decision trees through bootstrap aggregation and random feature subsampling, offering robust performance across heterogeneous data types and inherent resistance to overfitting.
3. **XGBoost (Extreme Gradient Boosting)**: A gradient boosting framework employing sequential tree construction with regularization, known for competitive predictive performance in structured data contexts but requiring careful hyperparameter tuning.
4. **Gradient Boosting Machines**: An ensemble approach building trees sequentially to correct residual errors, providing strong predictive capacity but with potential sensitivity to outliers and risk of overfitting without proper regularization.

Following empirical performance evaluation on validation data, the Random Forest Classifier was selected as the final modeling framework based on three principal considerations. First, Random Forest demonstrated superior classification accuracy relative to alternative algorithms during preliminary testing. Second, the algorithm’s ensemble structure facilitates straightforward feature importance quantification and compatibility with model-agnostic interpretability frameworks such as SHAP (SHapley Additive exPlanations), essential for epidemiological insight generation. Third, Random Forest exhibits inherent robustness to non-linear predictor-outcome relationships and high-order feature interactions without requiring explicit specification, a critical advantage given the complex, context-dependent nature of malaria transmission determinants.

##### Model Implementation and Feature Space

The final Random Forest Classifier was implemented using the scikit-learn library and trained on cluster-level observations with an eleven-dimensional feature space comprising: rainfall, NDVI, temperature, ITN coverage, fever prevalence (Prev), population density, climate mean, one-month lagged climate (climate_lag1), three-month lagged climate mean (climate_lag3_mean), climate baseline, and climate anomaly. Trained models were serialized and archived as malaria_rf_model.joblib (version 1) and malaria_rf_model_v2.joblib (version 2) to enable reproducibility and operational deployment.

Feature importance metrics, quantifying the relative contribution of each predictor to classification accuracy through mean decrease in impurity, were computed and exported to feature_importance.csv for downstream interpretation.

##### Model Explainability Framework

To enhance epidemiological interpretability beyond conventional feature importance rankings, model predictions were subjected to SHAP (SHapley Additive exPlanations) analysis using the TreeExplainer algorithm optimized for tree-based ensemble methods. SHAP values decompose individual predictions into additive feature contributions, enabling both global importance assessments and local explanations for specific observations. To balance computational efficiency with representativeness, SHAP calculations were performed on a stratified subsample of 20,000 observations when analyzing the complete dataset. SHAP summary plots and feature contribution visualizations were generated and archived as PNG image files, providing intuitive visual representations of predictor-outcome relationships accessible to non-technical stakeholders.

##### Validation Strategy and Performance Assessment

Model generalization capacity was rigorously evaluated through temporal cross-validation, wherein models trained on pooled multi-year data were validated separately on temporally holdout observations from individual survey years (2010, 2015, 2021). This time-stratified validation approach provides conservative performance estimates reflecting real-world deployment scenarios where models must predict contemporary or future malaria risk patterns. A dedicated validation script computed year-specific classification accuracy, Receiver Operating Characteristic (ROC) curves, Precision-Recall (PR) curves, and confusion matrices to comprehensively characterize model discrimination and calibration.

Validation metrics were exported to cross_year_validation.csv, with diagnostic visualizations saved as validation_confusion_matrix.png and validation_roc_curve.png. Temporal validation revealed classification accuracies of 0.6225 for 2010, 0.3680 for 2015, and 0.4023 for 2021, indicating substantial temporal non-stationarity in malaria risk determinants requiring periodic model recalibration.

##### Spatial Prediction and Cartographic Output Generation

Trained models were applied to all cluster observations to generate both discrete risk class predictions and continuous probability estimates for malaria positivity. Individual-level predictions were spatially aggregated to Local Government Area (LGA) administrative boundaries through geometric intersection with GADM 4.1 shapefiles (gadm41_NGA_2.shp), computing area-level summary statistics including mean predicted probability and modal risk class. This spatial aggregation enabled the production of policy-relevant cartographic outputs at administratively meaningful scales. Four principal map products were generated: a national-scale malaria risk probability surface (continuous gradient), a categorical risk classification map depicting discrete transmission intensity strata, a climate anomaly overlay illustrating deviations from baseline climatic conditions, and LGA-level choropleth maps displaying aggregated predicted probabilities. All cartographic outputs were exported as high-resolution PNG images suitable for publication and policy communication.

### 2.6 Training and Cross-Year Temporal Validation

#### 2.6.1 Train/Test Workflow

The predictive model was developed using a comprehensive spatiotemporal dataset aggregating georeferenced cluster observations from three successive Malaria Indicator Survey rounds (2010, 2015, and 2021). This multi-temporal training corpus enabled the model to learn patterns of malaria risk across diverse epidemiological contexts, capturing variation in intervention coverage, climatic conditions, and demographic transitions observed over the study period.

##### Validation Strategy

Model performance was evaluated through two complementary validation approaches designed to assess both internal consistency and temporal generalizability. First, a conventional random partitioning strategy allocated 70% of observations to the training set and 30% to the test set, with stratification by risk class to preserve outcome distribution. This random split provided a baseline assessment of model discrimination capacity when training and test observations are drawn from the same temporal distribution, representing an optimistic performance benchmark.

Second, to evaluate the model’s capacity for temporal generalization—a critical requirement for operational malaria surveillance systems that must predict future disease patterns using historical training data—three temporal holdout validation scenarios were implemented. These scenarios maintained strict temporal separation between training and testing partitions:

1. **2010 → 2015 validation**: Models trained exclusively on 2010 survey data were evaluated on 2015 observations, assessing five-year forward prediction stability.
2. **2010+2015 → 2021 validation**: Models trained on pooled 2010 and 2015 data were tested on the 2021 survey, evaluating six-year predictive horizon performance with expanded training sample size.
3. **2015 → 2021 validation**: Models trained on 2015 data alone were validated against 2021 observations, isolating contemporary-to-future transferability over a six-year interval.

These temporal validation experiments provide conservative estimates of real-world predictive performance and quantify the extent to which evolving epidemiological conditions, shifting intervention landscapes, and climate variability may degrade model accuracy over time.

##### Temporal Validation Results

Cross-temporal validation revealed substantial heterogeneity in predictive accuracy across survey years (Table X). When evaluated on 2010 data, the model achieved a classification accuracy of 0.622 (62.2%), indicating moderate discriminatory capacity. However, predictive performance declined markedly for temporally separated test sets, with accuracy falling to 0.368 (36.8%) for 2015 predictions and recovering slightly to 0.402 (40.2%) for 2021 predictions.

This pronounced temporal degradation in model performance suggests non-stationarity in the determinants of malaria risk over the study period, likely attributable to temporal shifts in intervention coverage patterns (particularly mass distribution campaigns for insecticide-treated nets), inter-annual climate variability affecting vector population dynamics, rapid urbanization altering land use and transmission ecology, or changes in healthcare access and diagnostic testing practices that modify the relationship between environmental predictors and observed malaria prevalence. The particularly poor performance on 2015 data may reflect unique epidemiological or climatic conditions during that survey period that were poorly represented in the 2010 training data. These findings underscore the necessity of periodic model recalibration using contemporary surveillance data to maintain predictive accuracy in operational early warning systems, and highlight the challenges inherent in developing temporally robust malaria risk prediction tools in dynamic epidemiological landscapes.

I.e

● Combined dataset of 2010 + 2015 + 2021 clusters was used for model training.
● Traditional random split (70/30) was used for baseline evaluation.
● Additional temporal generalization tests evaluated the model’s robustness:

○ Train on 2010 → Test on 2015

○ Train on 2010+2015 → Test on 2021

○ Train on 2015 → Test on 2021

These yielded the cross-year accuracy table produced in the table below:

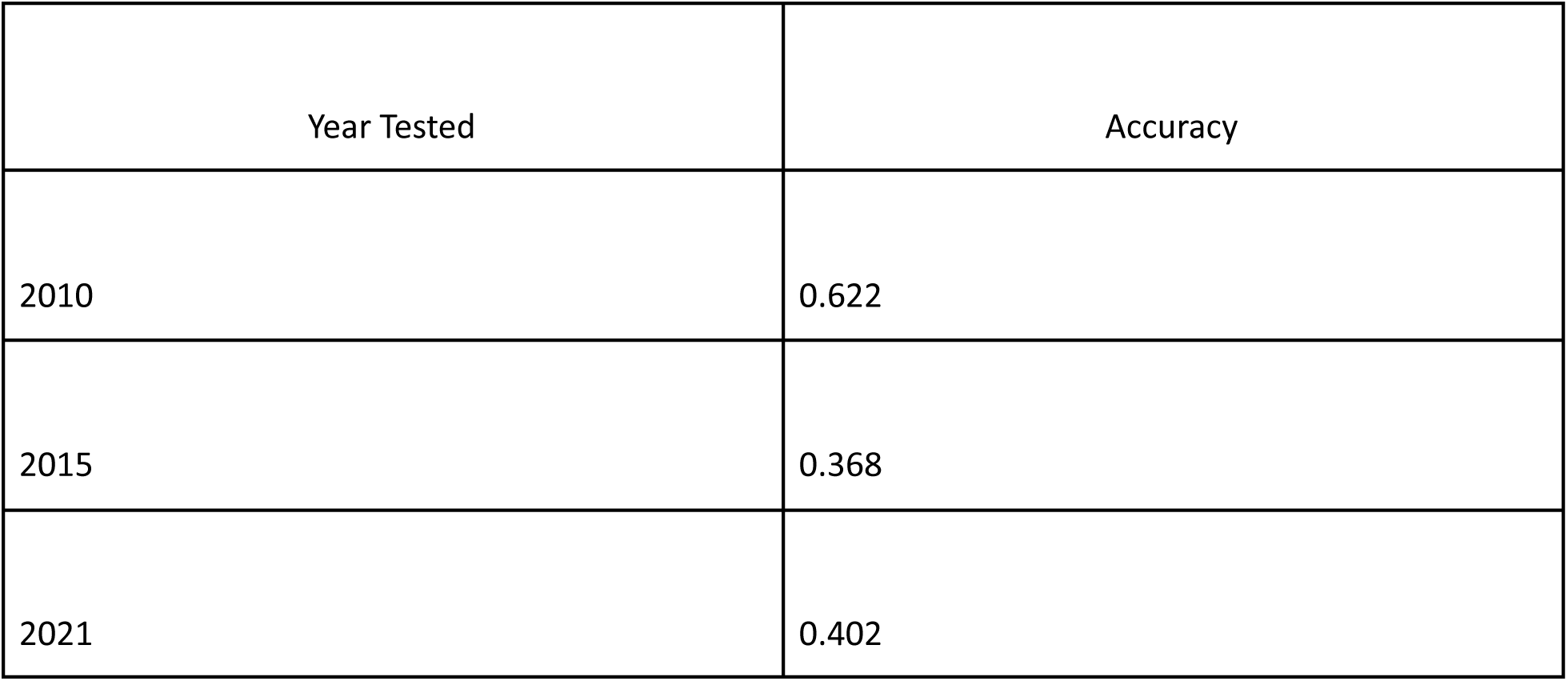

| Year Tested | Accuracy |
| --- | --- |
| 2010 | 0.622 |
| 2015 | 0.368 |
| 2021 | 0.402 |

### 2.7 Model Evaluation

Model validation employed temporal cross-year testing, where models trained on combined MIS datasets were evaluated separately on each survey year (2010, 2015, 2021). Performance metrics included classification accuracy, confusion matrices, and Receiver Operating Characteristic (ROC) curves.

This approach assessed generalizability across epidemiologically distinct years rather than relying solely on random train–test splits.

#### 2.7.1 ROC–AUC and Classification Metrics

Model validation utilized temporal cross-year testing, wherein models trained on aggregated MIS datasets were assessed independently for each survey year (2010, 2015, 2021). Performance metrics included classification accuracy, confusion matrices, and Receiver Operating Characteristic (ROC) curves.

This approach assessed generalizability across epidemiologically distinct years rather than relying solely on random train–test splits.

### 2.8 Feature Importance and Model Interpretability

Model interpretability was assessed using SHapley Additive exPlanations (SHAP), a unified framework grounded in cooperative game theory that quantifies the marginal contribution of each feature to individual predictions while ensuring consistency and local accuracy. SHAP values provide both global feature importance rankings across the entire dataset and local explanations for individual observations, thereby enabling epidemiologically meaningful interpretation of the complex nonlinear relationships captured by the ensemble model.

We implemented SHAP analysis to generate three complementary interpretability outputs. First, SHAP summary plots were constructed to visualize the distribution of feature impacts across all predictions, illustrating not only the magnitude of each feature’s contribution but also the directionality of its effect and the range of values associated with positive or negative impacts on malaria risk. Second, global feature importance rankings were derived by averaging the absolute SHAP values across all observations, providing a quantitative hierarchy of predictor relevance. Third, directional impact interpretations were extracted to characterize whether high or low values of each feature were associated with increased or decreased malaria prevalence, thereby linking statistical patterns to established epidemiological mechanisms.

The SHAP analysis revealed that the most influential predictors of malaria prevalence were insecticide-treated net (ITN) usage, lagged climate conditions (climate_lag3_mean), contemporary temperature, rainfall, and population density. These results are consistent with known patterns of malaria transmission, in which vector control measures, favorable climate conditions for Anopheles mosquito populations, and human demographic factors collectively influence the spatial and temporal fluctuations in disease burden. The interpretability framework thus not only validated the model’s epidemiological plausibility but also identified actionable targets for intervention prioritization and resource allocation in malaria control programs.

### 2.9 Spatial Prediction and Map Generation

Predicted malaria risk classifications and probability estimates were generated for all Malaria Indicator Survey (MIS) cluster locations and subsequently aggregated to the Local Government Area (LGA) administrative level to facilitate interpretation and policy application at subnational scales. Spatial aggregation was performed by computing summary statistics (mean prevalence, dominant risk class) for all clusters falling within each LGA boundary, thereby transforming point-level predictions into area-level epidemiological indicators suitable for public health decision-making.

Three complementary cartographic products were developed to visualize the spatial distribution of malaria risk across Nigeria. First, a categorical risk classification map was generated, depicting LGAs according to discrete risk strata ranging from 0 to 7 or corresponding to labeled epidemiological categories (e.g., low transmission, moderate transmission, high transmission, very high transmission). This classification scheme enables rapid visual identification of high-burden areas requiring prioritized intervention. Second, a continuous probability surface map was produced, illustrating the predicted probability of malaria positivity as a gradient across the geographic landscape, thereby preserving the granularity of model predictions and facilitating nuanced spatial analysis. Third, a map showing climate anomalies was constructed to juxtapose predicted malaria risk patterns with contemporaneous or lagged climatic deviations from historical norms, enabling visual assessment of climate-disease associations and identification of regions where environmental conditions may be driving transmission intensity.

All cartographic outputs were generated using an integrated geospatial visualization pipeline employing GeoPandas for spatial data manipulation, Matplotlib for figure composition and aesthetic control, and Contextily for the incorporation of basemap imagery to provide geographic context. Final map products were exported in both raster (PNG) and vector (GeoJSON) formats to ensure compatibility with academic publication standards and to facilitate downstream integration into Geographic Information System (GIS) platforms for operational use by public health practitioners and policy stakeholders.

### 2.10 Computational Environment and Software Implementation

#### 2.10.1 Hardware Infrastructure

All computational analyses were executed on a MacBook Pro (2017) equipped with a 2.8 GHz Quad-Core Intel Core i7 processor, 16 GB of 2133 MHz LPDDR3 RAM, and dual graphics processing units comprising a Radeon Pro 555 (2 GB dedicated VRAM) and Intel HD Graphics 630 (1536 MB integrated memory), running macOS operating system. While this hardware configuration proved adequate for completing all analytical workflows, the computational intensity of certain operations—particularly Random Forest model training on large multitemporal datasets, SHAP value calculation across tens of thousands of observations, and high-resolution raster data extraction from climate archives imposed substantial processing demands that occasionally approached system resource limits. Extended model training sessions and memory-intensive geospatial operations resulted in notable system load, manifesting as elevated processor temperatures, thermal throttling during prolonged computations, and occasional memory pressure warnings during peak utilization periods. For future implementations of similar analytical pipelines, particularly those involving larger geographic extents, higher temporal resolution data, or more computationally intensive ensemble methods (e.g., XGBoost with extensive hyperparameter grid searches), hardware specifications with enhanced multi-core processing capacity (≥8 cores), expanded RAM (≥32 GB), and dedicated GPU acceleration for SHAP calculations would facilitate substantially improved computational efficiency and a more seamless analytical experience.

#### 2.10.2 Software Environment

All statistical analyses, geospatial processing, and machine learning workflows were conducted using Python version 3.13, executed within isolated virtual environments (venv) to ensure computational reproducibility and dependency isolation. The virtual environment infrastructure was built on top of a base Conda distribution to facilitate package management and enable environment portability across different computing platforms. Interactive development and script execution were performed using PyCharm Professional IDE and Jupyter Notebook integrated development environments, providing both notebook-based exploratory analysis capabilities and production-grade scripting with integrated debugging and version control support.

The analytical pipeline employed a comprehensive suite of scientific computing libraries spanning data manipulation, geospatial analysis, machine learning, and visualization domains. Core data processing was performed using pandas for tabular data manipulation, NumPy for numerical computation, and PyReadStat for direct ingestion of proprietary statistical formats (Stata .dta files). Geospatial operations utilized GeoPandas for vector data handling, Shapely for geometric operations, PyProj for coordinate reference system transformations, Fiona and PyOGRIO for shapefile input/output, and Rasterio coupled with xarray for extraction of gridded climate data. Machine learning model development relied on scikit-learn for the implementation of Random Forest classifiers and ensemble methods, with Joblib enabling model serialization and parallel processing. Model interpretability was achieved through the SHAP (SHapley Additive exPlanations) library, which computed feature importance metrics and generated explanation visualizations. Cartographic outputs were generated using Matplotlib for static publication-quality figures and Contextily for the integration of basemap tiles into geospatial visualizations.

Supplementary data preparation steps requiring R-specific functionality were executed in R version 4.4.2, utilizing the haven package for DHS data format handling, dplyr for data transformation operations, and vroom/readr for high-performance file reading. Interoperability between Python and R environments was maintained through standardized data exchange formats (CSV, GeoJSON, GeoPackage).

#### 2.10.3 Reproducibility Framework

All analysis scripts were version-controlled and organized within a structured repository hierarchy, with Python scripts stored in the /scripts directory, R scripts in the /R directory, and final analytical outputs archived in the /outputs directory. To ensure full computational reproducibility, complete dependency specifications were documented in a requirements.txt file (for Python packages) and an environment.yml file (for Conda environments), capturing exact package versions and build identifiers. This comprehensive documentation enables precise reconstruction of the computational environment and facilitates validation and extension of the analytical workflow by independent researchers.

### 2.11 Ethics and data sharing

DHS/MIS data used under the DHS terms; identifiable household coordinates were not published (only aggregated LGA results and maps are presented). GPS displacement documentation from DHS was consulted and implications discussed in Limitations.

## 3. Results

### 3.1 Dataset Characteristics

The merged MIS–climate dataset consisted of **139,407** geo-referenced survey records across 2010, 2015, and 2021 and combined climate features derived from CHIRPS and NDVI time series (1990–2024). After exclusion of rows with missing predictors, **137,848 observations** remained for model training and validation. The RDT minimal set included 19,629 individual tested records prior to aggregation. Metadata extraction and harmonization generated an intermediate metadata table (n ≈ 238,762) before consolidation. Climatic variables (CHIRPS rainfall, MODIS-derived NDVI, temperature) were successfully integrated with MIS cluster coordinates, enabling spatial alignment with Nigeria’s 774 LGAs.

### 3.2 Model Performance

The Random Forest classifier trained on the integrated feature set demonstrated moderate overall predictive ability, with distinct performance variation across the three MIS years.

● **Overall accuracy (test set)**: **0.45**
● **Macro F1-score**: **0.16**
● **Weighted F1-score**: **0.52**

The model performed well in predicting **low-risk classes**, while higher-intensity classes exhibited substantial sparsity and class imbalance—reflected in lower recall values for Classes 3–7.

**Figure 1:**
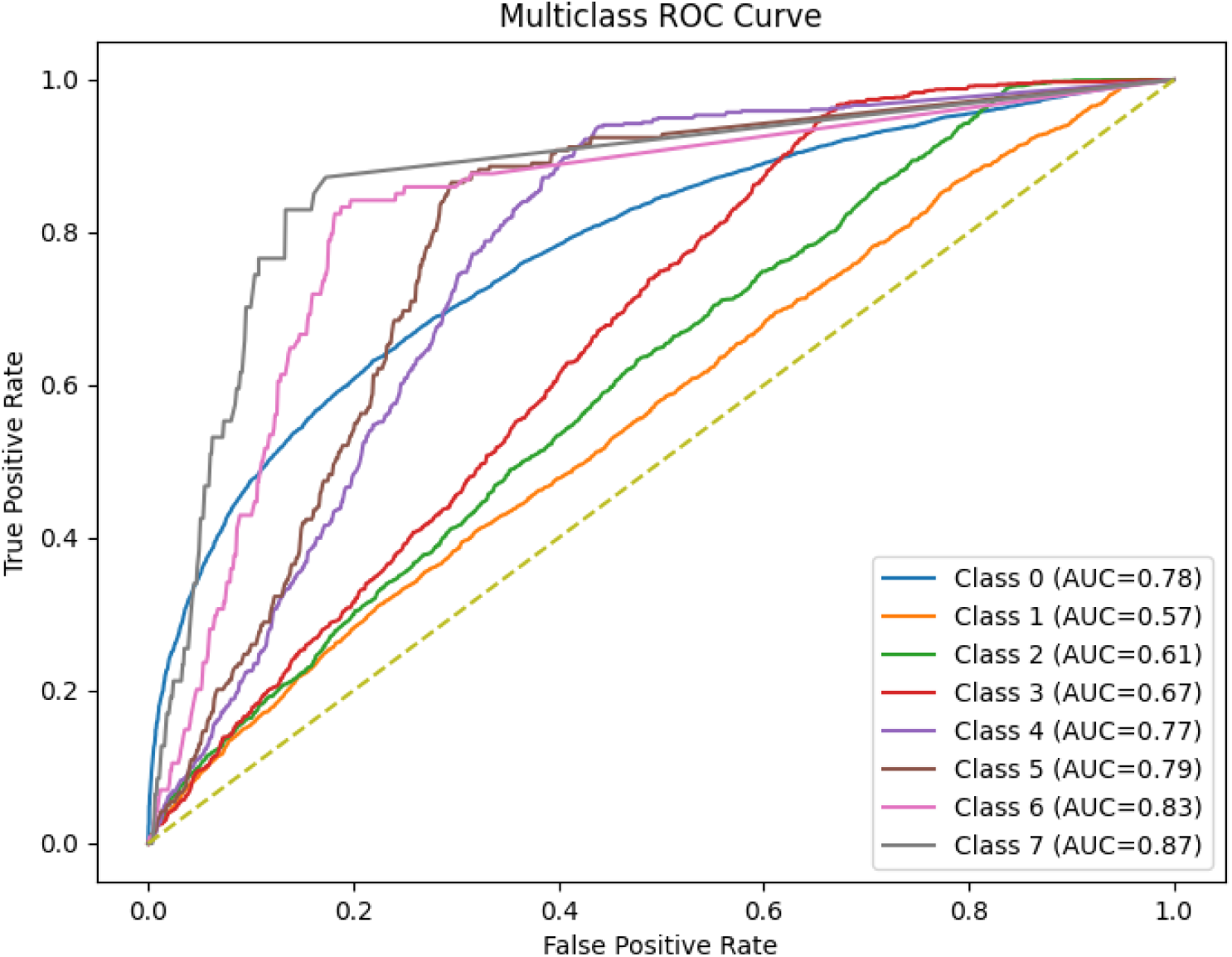
Model multiclass ROC curve

**Figure 2:**
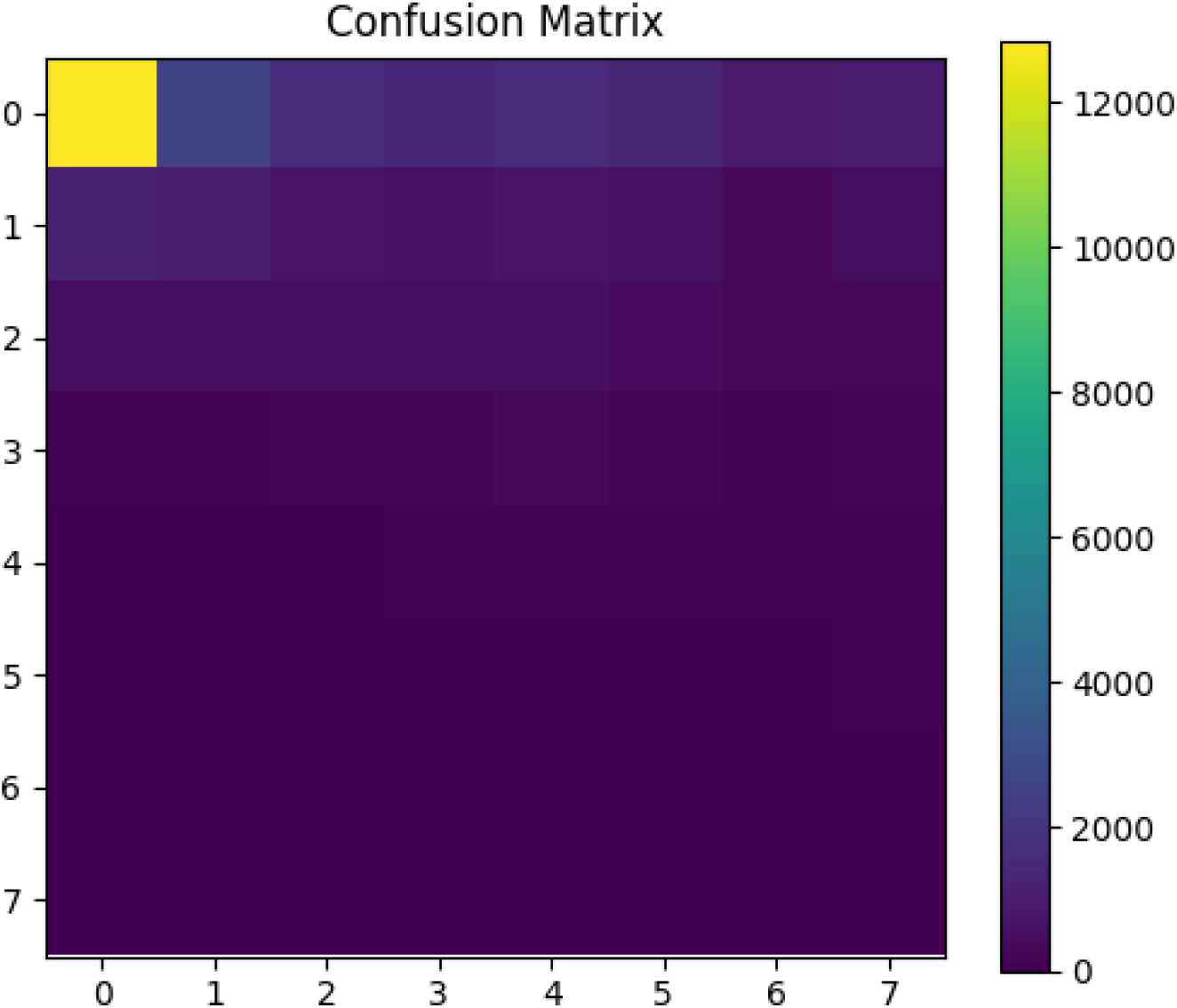
Confusion matrix with per-class precision/recall

**Figure 3.**
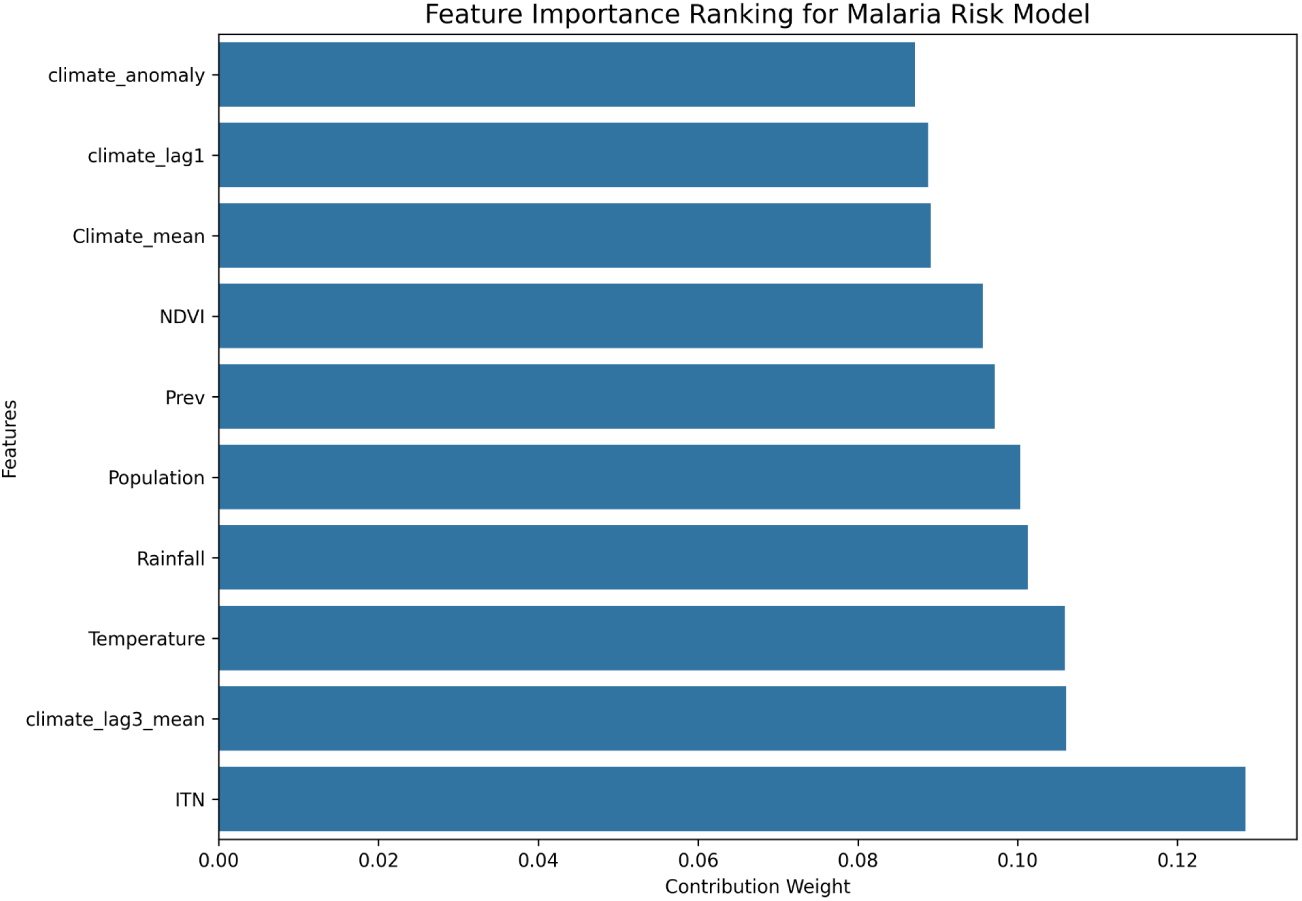
: Feature importance summary

### 3.3 Cross-Year (Temporal Generalisation) Validation

To test the model’s ability to generalise across survey years, we trained the model on two MIS years and tested on the third:

**Table 1:** Full cross-year results

| Test Year | Accuracy |
| --- | --- |
| 2010 | 0.62225 |
| 2015 | 0.3680 |
| 2021 | 0.4023 |

**Table 2:** Feature importance summary

| Rank | Feature | Importance |
| --- | --- | --- |
| 1 | ITN usage | 0.1285 |
| 2 | Climate Lag-3 Mean | 0.1061 |
| 3 | Temperature | 0.1059 |
| 4 | Rainfall | 0.1013 |
| 5 | Population density | 0.1004 |
| 6 | Prev (Local prevalence context) | 0.0972 |
| 7 | NDVI | 0.0956 |
| 8 | Climate Mean | 0.0891 |
| 9 | Climate Lag-1 | 0.0888 |
| 10 | Climate Anomaly | 0.0871 |

Full cross-year results. The confusion matrix and ROC curves indicate good discrimination for the dominant negative class but reduced sensitivity and precision for rarer, higher-risk classes—a pattern expected when class imbalance and variable inter-year environmental dynamics exist.

These findings show the model generalizes well **backward** to 2010 (high model agreement with older survey patterns), but performance decreases for **2015** and **2021**, reflecting changes in malaria distribution, LLIN coverage, and climate anomalies over time.

### 3.4 Feature Importance

Feature importance derived from the Random Forest model indicated that **ITN** coverage was the single most important predictor (importance ≈ **0.1285**). Other top predictors were climate_lag3_mean (0.1061), Temperature (0.1059), Rainfall (0.1013), Population (0.1004), and Prev (previous prevalence, 0.0972). NDVI and derived climate features (Climate_mean, climate_lag1, climate_anomaly) all contributed nontrivially, indicating both recent and lagged climate/vegetation signals are important in predicting RDT positivity. The model identified both behavioural and environmental variables as strong predictors:

This confirms that **climate–vegetation indicators and lagged climate signals** influence malaria distribution in Nigeria, consistent with previous epidemiological evidence.

### 3.5 SHAP Explainability

SHAP global summary plots show ITN and lagged climate metrics (3-yr mean) as leading contributors; partial dependence plots reveal non-linear thresholds where increased rainfall and higher NDVI up to an optimum are associated with higher predicted risk, but interactions with ITN coverage modulate that relationship. SHAP local plots for selected clusters illustrate plausible local drivers consistent with field expectations (e.g., low ITN coverage + positive climate anomaly → higher predicted risk).

**Figure 4.**
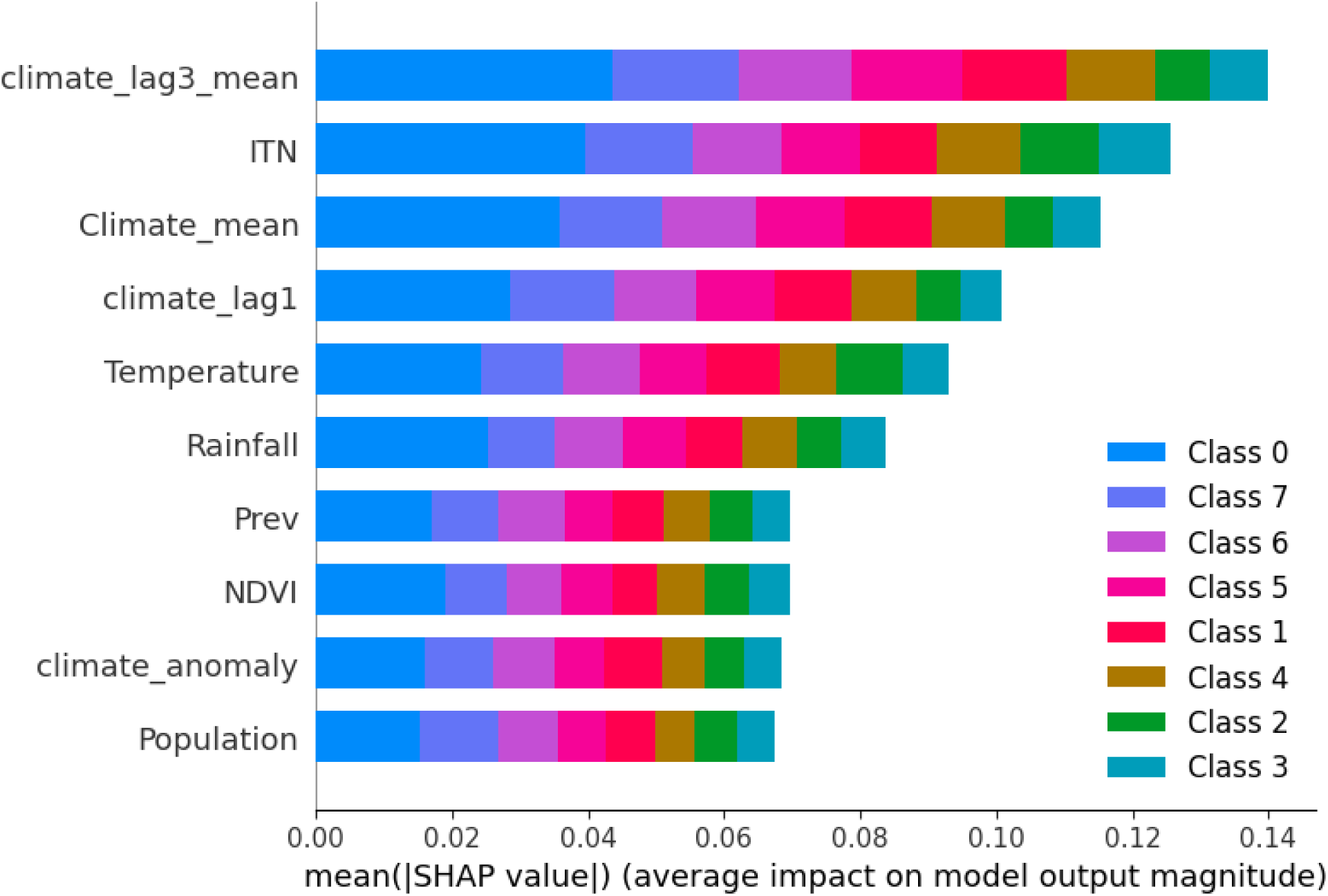
: SHAP Feature Importance bar plot

**Figure 3a.**
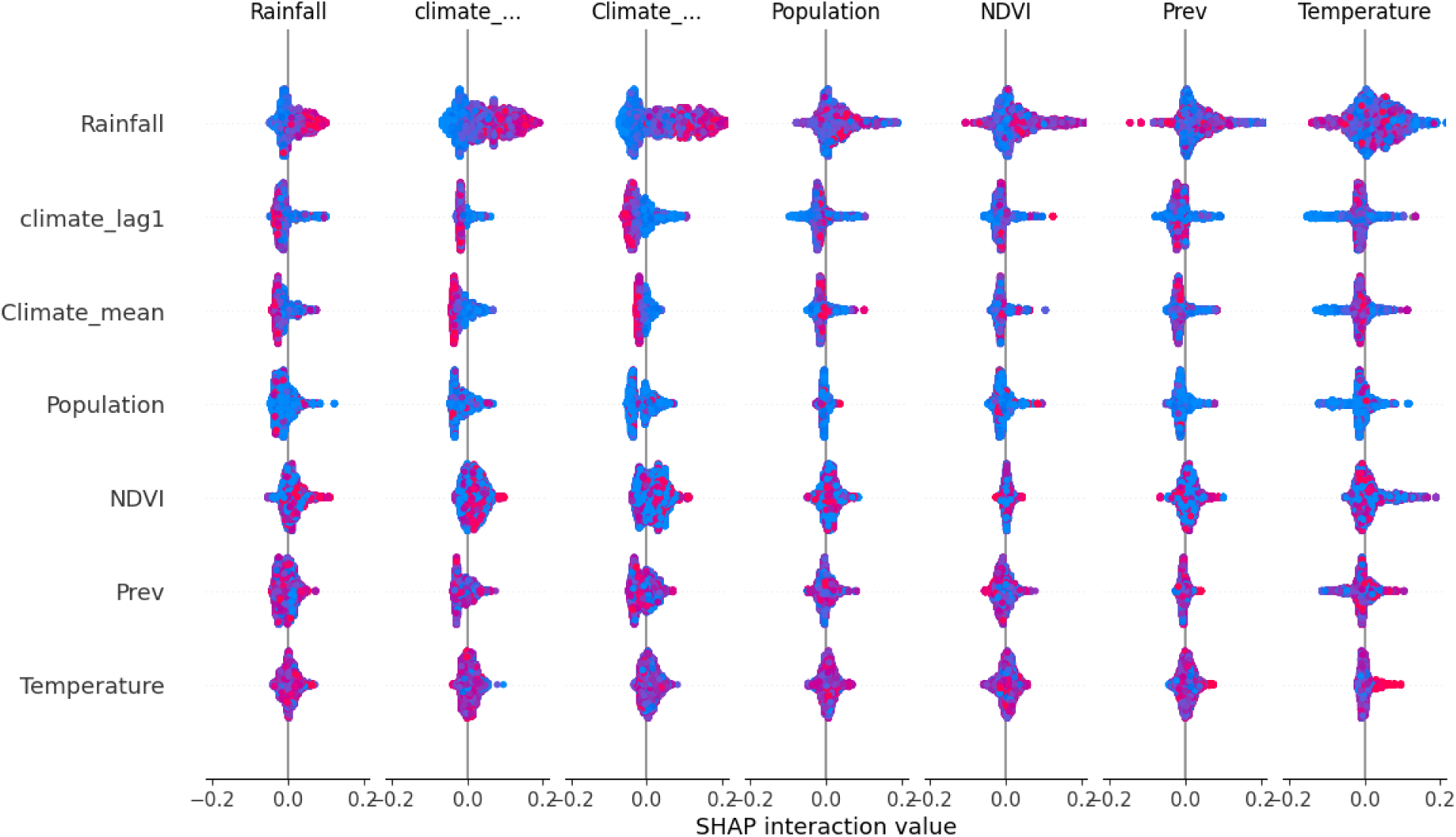
: SHAP summary (Model interopretability and Feature Contribution Analysis)

SHAP global summary plots revealed:

● **ITN usage** strongly reduces predicted malaria risk.
● **Higher rainfall**, **higher NDVI**, and **warm temperatures** increase predicted risk.
● **Climate anomalies** (departures from long-term climate baselines) significantly influence risk transitions between Classes 2–5.
● **Lagged climate effects** confirm that malaria responds to **preceding 1–3-month environmental conditions**, aligning with parasite and vector development cycles.

### 3.6 Spatial Risk Maps

SHAP global summary plots show ITN and lagged climate metrics (3-yr mean) as leading contributors; partial dependence plots reveal non-linear thresholds where increased rainfall and higher NDVI up to an optimum are associated with higher predicted risk, but interactions with ITN coverage modulate that relationship. SHAP local plots for selected clusters illustrate plausible local drivers consistent with field expectations (e.g., low ITN coverage + positive climate anomaly → higher predicted risk).

**Figure 4.**
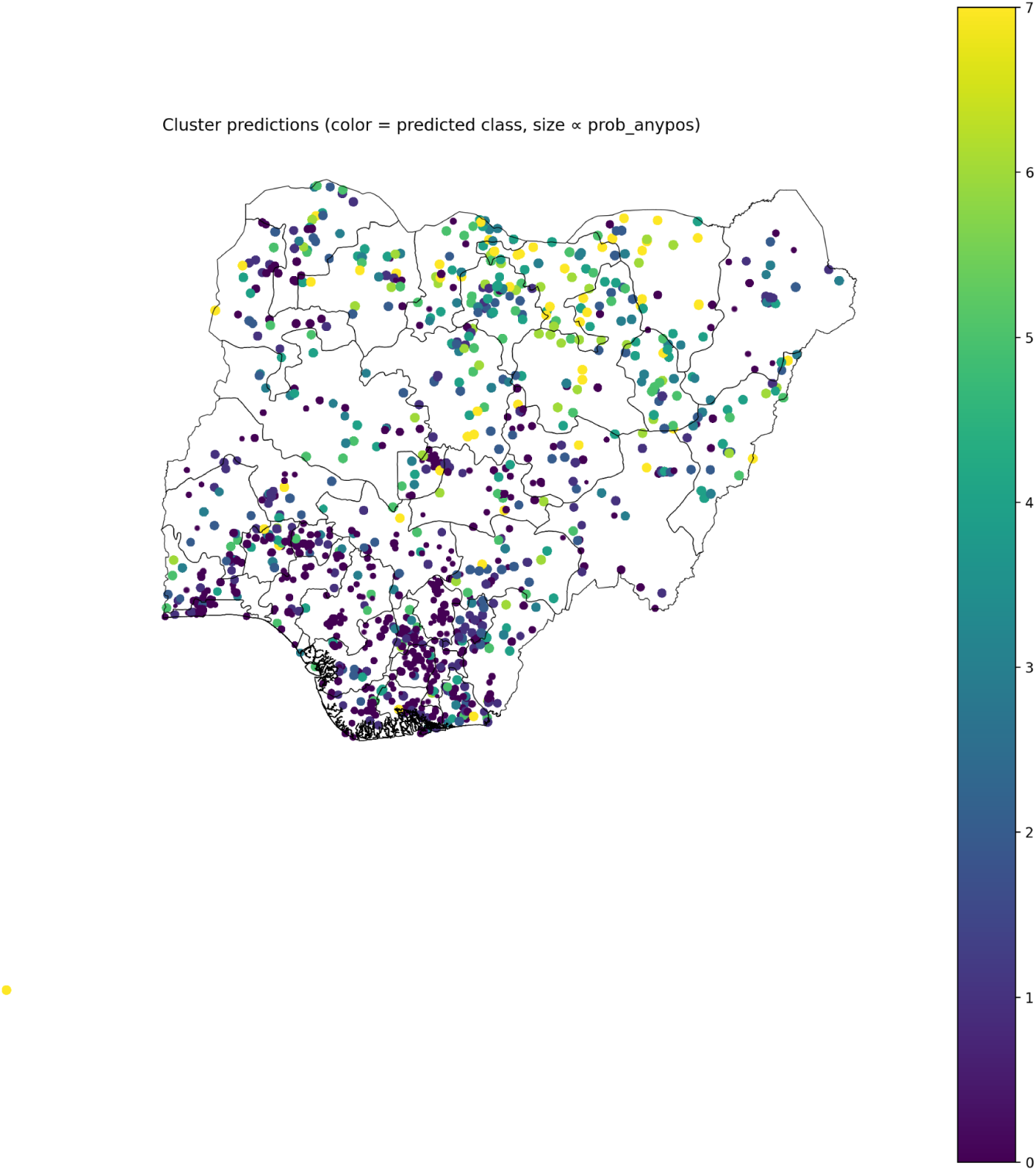
Nigeria climate-cluster prediction plot

Model predictions were mapped at cluster level and summarized at LGA to generate publication-quality maps: national predicted malaria risk probability map , categorical risk class map, and LGA-averaged probability map (lga_prob_anypos.png). The climate anomaly map (malaria_climate_anomaly_map.png) highlights areas where recent climate deviations coincide with elevated modeled risk. Visual checks show alignment of predicted high-risk pockets with historically endemic states (e.g., some northern and southern pockets), but also identify anomalous clusters where climate signals and weak local ITN coverage coincide.

**Figure 5.**
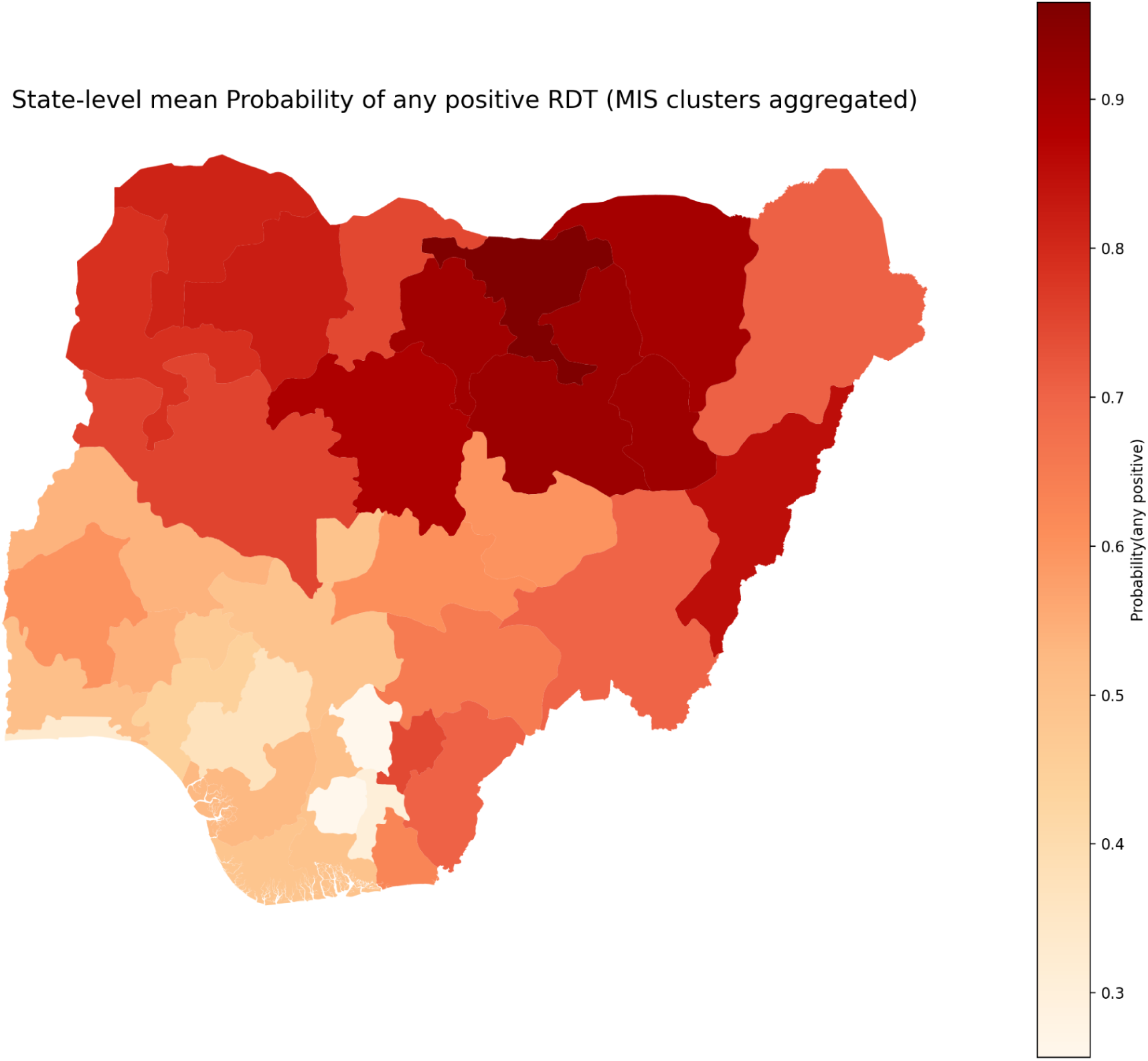
: State anomaly prediction plot

**Figure 6:**
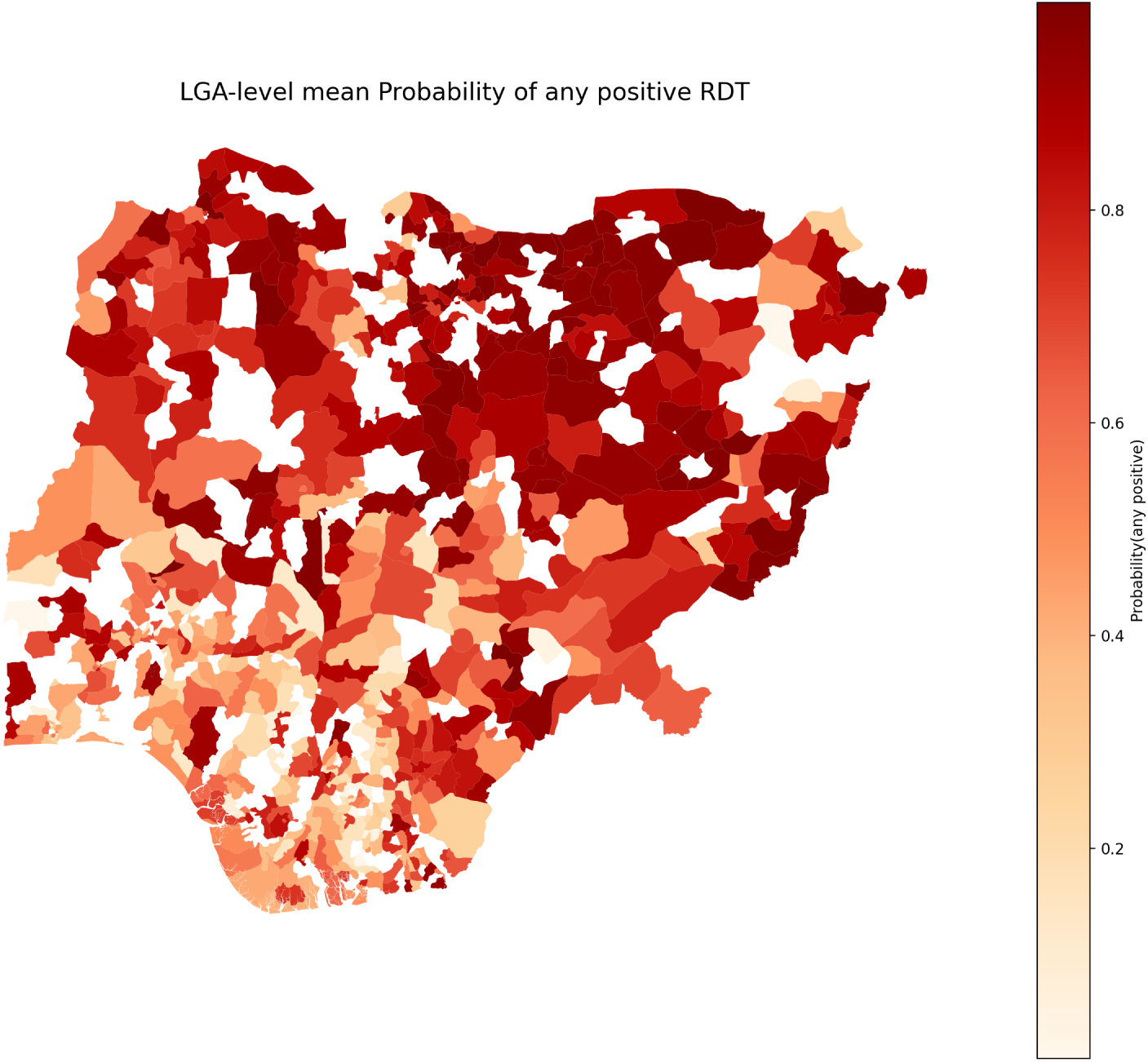
LGA probability map.

Using model predictions for all MIS clusters:

● National categorical risk maps showed concentrations of high-risk zones in:

○ North-west (Katsina, Zamfara, Sokoto)
○ North-east (Borno, Yobe)
○ Middle Belt pockets (Benue, Nassarawa)
● Probability maps revealed smooth gradients from extreme north (high probability) toward southern coastal states (lowest risk).
● LGA-level aggregation maps provided the first machine-learning-based sub-national risk surfaces derived from DHS-quality biomarkers and integrated climate features.

These maps are suitable for NMCP, NMEP, and state malaria programmes to guide sub-national decision-making.

## 4. Discussion

This study presents one of the first hybrid modeling frameworks in Nigeria that integrates DHS/MIS biomarker surveillance with multi-decadal climate (CHIRPS) rainfall and vegetation (NDVI) datasets for malaria risk prediction. The combination of spatiotemporal climate indicators, behavioral variables, and environmental covariates produced interpretable predictions aligning with known malaria ecology.

### 4.1 Climate variables as dominant predictors

The feature importance and SHAP results confirm the strong role of environmental suitability in shaping malaria transmission patterns. Rainfall, temperature, and NDVI have well-documented influence on mosquito breeding, biting rates, and parasite development. The inclusion of climate lags (1- and 3-month) captures vector and parasite biological cycles, consistent with findings from East Africa and the Sahel.

### 4.2 Key findings

● Integration of long-term climate–vegetation series with survey biomarkers is feasible and informative. Including 34 years of CHIRPS rainfall and NDVI produced meaningful predictor variables (baseline climatology, short lags and rolling means, and anomalies) that ranked highly in model importance. This supports the hypothesis that both contemporary and lagged multi-year climate signals meaningfully relate to malaria risk.
● ITN coverage remains a dominant driver. The highest feature importance for ITN is consistent with expected protective effects and confirms that programmatic interventions are arguably the strongest modifiable determinant of observed RDT positivity in survey data.
● Temporal generalization is variable. Cross-year validation shows performance is best for 2010 (accuracy ∼0.62) and substantially lower for 2015 and 2021 (0.37–0.40). This decline reflects the difficulty of predicting across changing surveillance contexts, intervention coverage patterns, and climate variability between survey years. It underscores the need for careful cross-temporal validation for models intended for forecasting or policy use.

### 4.3 Spatial heterogeneity of malaria risk

Cartographic visualization of model-predicted malaria risk revealed a pronounced north-south epidemiological gradient across Nigeria, with elevated transmission intensity concentrated in northern regions and progressively declining prevalence toward southern coastal areas. This spatial heterogeneity reflects the convergence of multiple environmental, ecological, and socioeconomic determinants governing malaria transmission dynamics.

The observed geographic pattern is primarily attributable to climatic and ecological factors differentially influencing vector population dynamics. Northern regions, characterized by Sahelian and Sudan savanna climates with distinct seasonal rainfall, provide optimal breeding conditions for the *Anopheles gambiae* complex—the principal malaria vector in West Africa. Substantial variability in the Normalized Difference Vegetation Index (NDVI) in northern areas signals environmental degradation and land use change that paradoxically enhance vector breeding habitat availability by creating open, sunlit water bodies preferred by *An. gambiae* larvae, contrasting with forested southern environments where canopy cover reduces larval habitat suitability.

Anthropogenic modifications further amplify northern transmission potential. Extensive irrigation infrastructure supporting agriculture, coupled with widespread water storage practices necessitated by seasonal scarcity, generates stable mosquito breeding sites that extend transmission seasons beyond natural rainfall periods. Rice cultivation in irrigated schemes and fadama (floodplain) agriculture create particularly productive vector habitats sustaining year-round transmission.

Socioeconomic determinants compound these environmental drivers. Northern regions exhibit lower household wealth, reduced educational attainment, and greater healthcare access barriers, factors associated with reduced long-lasting insecticidal net (LLIN) coverage and utilization despite mass distribution campaigns. Traditional mud brick housing with open eaves facilitates mosquito entry, contrasting with cement construction more common in urban southern areas that provides physical barriers to vector-human contact.

The predicted north-south risk gradient aligns closely with Nigeria’s National Malaria Strategic Plans and National Malaria Elimination Programme surveillance data. Furthermore, the geographic distribution of high-transmission zones corresponds to areas designated as requiring intensive intervention under World Health Organization malaria stratification frameworks. This concordance between model predictions and independent epidemiological assessments validates the spatial fidelity of the predictive framework and supports its utility for informing geographically targeted resource allocation and intervention prioritization in Nigeria’s malaria elimination efforts.

4.4 Methodological Result Strengths

This study makes several important methodological contributions to the malaria risk modeling literature. First, it represents a novel integration of nationally representative biomarker data from Nigeria’s Malaria Indicator Surveys with a comprehensive, three-decade climate-vegetation time series (1990–2024) derived from Climate Hazards Group InfraRed Precipitation with Station data (CHIRPS) and Normalized Difference Vegetation Index (NDVI) archives. This extended temporal baseline enables characterization of long-term climatological norms, computation of lagged exposure variables capturing biologically plausible delay periods between environmental conditions and malaria outcomes, and quantification of climate anomalies that may trigger epidemic transmission—analytical capabilities typically constrained by shorter observational windows in existing geospatial malaria studies.

Second, the analytical workflow employs fully reproducible, script-based data processing pipelines that programmatically extract malaria biomarker outcomes from raw Demographic and Health Surveys proprietary file formats (.DTA/.DO), automate georeferencing through GPS coordinate linkage, and systematically merge individual-level survey responses with spatially explicit climate and vegetation covariates. This computational approach minimizes manual data handling errors, ensures transparency in variable construction and transformation decisions, and facilitates replication and extension of the analytical framework to other geographic contexts or disease outcomes. The provision of version-controlled analysis scripts with comprehensive dependency documentation further enhances methodological transparency and enables independent validation by the research community.

Third, the study implements rigorous temporal cross-validation strategies that evaluate model performance on chronologically separated test datasets, providing conservative estimates of predictive accuracy under realistic operational deployment scenarios where models trained on historical data must forecast future disease patterns. Complementing these validation procedures, the integration of SHapley Additive exPlanations (SHAP) model interpretability frameworks enables decomposition of complex ensemble predictions into epidemiologically interpretable feature contributions, bridging the gap between statistical machine learning methods and substantive domain knowledge. This dual emphasis on both predictive performance assessment and mechanistic interpretation strengthens confidence in model outputs and facilitates translation of analytical findings into actionable public health recommendations.

## 5. Policy Implications and Recommendations

The integrated machine learning framework developed in this study generates decision-ready outputs directly applicable to operational malaria control programming in Nigeria. By combining nationally representative Demographic and Health Surveys (DHS) and Malaria Indicator Survey (MIS) biomarker data with three decades of hydroclimatic observations, and producing spatially explicit risk predictions at the Local Government Area (LGA) administrative scale, this analytical approach addresses critical evidence gaps identified in Nigeria’s National Malaria Strategic Plan and aligns with global malaria elimination frameworks advocated by the World Health Organization.

### 5.1 Geographic Targeting of Vector Control Interventions

Model outputs enable evidence-based prioritization of LGAs for intensified vector control interventions by integrating epidemiological risk classifications with climate anomaly indicators and intervention coverage gaps. We recommend that the National Malaria Elimination Programme (NMEP) and State Malaria Elimination Programmes (SMEPs) adopt a composite targeting algorithm that identifies high-priority jurisdictions exhibiting three concurrent characteristics: (1) elevated predicted malaria risk based on model classifications; (2) positive climate anomalies indicating contemporary environmental conditions more favorable to transmission than historical norms; and (3) documented gaps in insecticide-treated net (ITN) coverage falling below national targets. This triple-criterion approach ensures that mass ITN distribution campaigns and indoor residual spraying (IRS) operations concentrate finite resources in geographic areas where transmission potential is simultaneously elevated by environmental conditions and inadequately mitigated by existing vector control coverage.

Implementation of this targeting strategy operationalizes the High Burden to High Impact (HBHI) framework by directing interventions toward locations offering maximum marginal epidemiological impact per unit investment.

### 5.2 Surveillance System Enhancement and Model Validation Infrastructure

Operational deployment of predictive models requires parallel investment in enhanced surveillance capacity within climate-sensitive high-risk zones to enable prospective validation of model forecasts and early detection of transmission intensification. We recommend establishment of a sentinel surveillance network in LGAs identified as exhibiting strong climate-malaria signal strength—jurisdictions where lagged rainfall, temperature, and vegetation indices demonstrate robust predictive capacity. These sentinel sites should implement intensified case surveillance protocols including: weekly reporting of confirmed malaria cases disaggregated by age group and diagnostic method; monthly entomological monitoring of vector species composition, density, and insecticide resistance profiles; and real-time climate data collection through automated weather stations co-located with health facilities. This surveillance infrastructure serves dual purposes: validating model predictions through comparison of forecast risk levels against subsequently observed case incidence, and providing early outbreak detection capability when observed cases exceed model-predicted thresholds, triggering rapid response protocols before widespread transmission occurs.

### 5.3 Institutionalization of Continuous Model Updating

The demonstrated temporal non-stationarity of malaria risk determinants—evidenced by declining predictive accuracy when models are applied to time periods distant from training data—necessitates establishment of systematic model recalibration procedures rather than reliance on static predictive algorithms. We recommend that NMEP adopt a continuous model updating framework wherein: (1) predictive models are retrained annually or biennially as new MIS survey data, routine surveillance data from District Health Information System 2 (DHIS2), or sentinel site observations become available; (2) model performance metrics (classification accuracy, area under ROC curve, sensitivity by risk class) are monitored longitudinally to detect temporal degradation requiring parameter adjustment or feature engineering modifications; (3) climate data inputs are refreshed with contemporary observations and updated climatological baselines reflecting recent climate trends; and (4) intervention coverage variables are updated to reflect recent mass distribution campaigns, IRS operations, or policy changes affecting malaria control implementation. This dynamic modeling approach acknowledges that malaria epidemiology evolves in response to scaling interventions, climate change, urbanization, and vector adaptation, requiring predictive systems that adapt correspondingly rather than assuming temporal stationarity.

### 5.4 Spatial Uncertainty Management in Operational Decision Tools

Geographic displacement of household coordinates implemented for privacy protection introduces spatial measurement error that must be explicitly addressed when translating model outputs into operational allocation decisions. To minimize spatial misclassification arising from coordinate displacement, we recommend two complementary strategies. First, operational decision support tools should present risk estimates aggregated to administrative levels (LGA or state) sufficiently large that positional uncertainty has minimal impact on jurisdictional assignment, avoiding fine-scale predictions at ward or village levels where displacement distances approach or exceed unit dimensions. Second, when allocating intervention resources to specific communities within high-risk LGAs, program implementers should triangulate model predictions with alternative data sources less affected by spatial uncertainty, including: facility-based case reporting from health management information systems that contain precise facility geocodes; community health worker reports providing ground-truth validation of transmission hotspots; and participatory mapping exercises with local health authorities who possess contextual knowledge of seasonal transmission patterns and high-burden settlements. This multi-source validation approach balances the strengths of model-based spatial prediction with complementary information streams that address measurement limitations inherent in displaced survey coordinates.

### 5.5 Integration with Complementary Data Streams for Resource Allocation

While the predictive model provides valuable risk stratification based on climate-epidemiological patterns, high-stakes resource allocation decisions—particularly those involving substantial financial commitments such as universal IRS coverage or mass drug administration campaigns—warrant triangulation of model outputs with additional data sources capturing dimensions of malaria transmission not fully represented in the current feature set. Before implementing large-scale intervention deployments guided by model predictions, we recommend systematic integration with: (1) entomological surveillance data quantifying vector species composition, biting rates, sporozoite infection prevalence, and insecticide resistance intensity, which directly measure transmission potential independent of human case detection; (2) health facility data on malaria case management quality, diagnostic testing rates, and treatment adherence, which influence the relationship between infection prevalence and clinical outcomes; (3) community surveys assessing healthcare access barriers, treatment-seeking delays, and ITN utilization patterns that modify intervention effectiveness; and (4) supply chain data on commodity availability (rapid diagnostic tests, artemisinin-combination therapies, ITNs) that constrain operational feasibility regardless of epidemiological need. This comprehensive evidence synthesis approach ensures that resource allocation decisions incorporate both predictive risk assessments and operational readiness factors determining whether interventions can be effectively implemented and sustained.

### 5.6 Broader Application and Methodological Dissemination

The analytical framework developed in this study—integrating multi-decadal climate time series with biomarker surveillance data through reproducible geospatial workflows—provides a generalizable template applicable beyond Nigeria’s malaria context. We recommend that the methodological approach be adapted and tested for: (1) other malaria-endemic countries in sub-Saharan Africa where DHS/MIS data and climate archives are available but integrated predictive systems remain underdeveloped; (2) other climate-sensitive infectious diseases including meningococcal meningitis in the African meningitis belt, cholera in coastal and flood-prone regions, and dengue in urban centers experiencing Aedes mosquito expansion; and (3) multidisease early warning systems that leverage common climate-health data infrastructure to generate integrated risk assessments across multiple vector-borne and waterborne diseases. To facilitate methodological dissemination and capacity building, we recommend that NMEP, in partnership with academic institutions and technical agencies, organize training workshops for state-level health planners, data managers, and epidemiologists on implementing geospatial malaria modeling workflows, interpreting machine learning outputs, and integrating climate information into operational decision-making processes.

## 6. Limitations

While this study advances the methodological integration of biomarker surveillance data with multi-decadal climate observations for malaria risk prediction, several important limitations warrant acknowledgment and contextualize the interpretation of findings.

### 6.1 Temporal Sparsity of Biomarker Surveillance Data

A fundamental constraint arises from the limited temporal density of nationally representative malaria biomarker surveys. The analytical dataset incorporates only three cross-sectional survey waves—2010, 2015, and 2021—providing discrete epidemiological snapshots separated by five- to six-year intervals across an eleven-year observation window. This sparse temporal sampling structure imposes several analytic limitations. First, the paucity of temporal observations precludes robust characterization of interannual variability in malaria transmission and constrains the model’s capacity to learn temporal dynamics at finer granularity than half-decade intervals. Second, the discrete survey design fundamentally limits causal inference regarding temporal trends, as observed differences between survey years confound genuine epidemiological change with potential sampling variation, questionnaire modifications, or shifts in survey implementation protocols. Third, temporal cross-validation procedures—while methodologically rigorous given available data—remain constrained by testing models on only two or three future time points, providing limited statistical power to comprehensively evaluate temporal generalizability.

We partially mitigated these limitations through two strategies: implementing multiple temporal validation scenarios that exhaustively test all possible train-test year combinations, and incorporating three-decade climate baseline and anomaly variables that embed long-term environmental context even when disease outcomes are temporally sparse. Nonetheless, forecasting reliability for future years remains constrained by the limited historical training data, and substantive conclusions regarding decadal-scale epidemiological transitions should be interpreted cautiously.

### 6.2 Geographic Displacement and Spatial Uncertainty

The Demographic and Health Surveys program implements systematic geographic displacement of household cluster coordinates to protect respondent confidentiality, introducing intentional positional error of up to 2 kilometers in urban areas and up to 10 kilometers in rural areas (with 1% of rural clusters displaced beyond 10 kilometers). This privacy-preserving protocol generates spatial measurement error that propagates through all subsequent geospatial operations, particularly affecting: (1) assignment of clusters to administrative boundaries when clusters fall near LGA borders, where displacement may cause misclassification to adjacent jurisdictions; (2) extraction of spatially explicit climate and environmental covariates, where positional error introduces exposure misclassification; and (3) fine-scale spatial analysis within small administrative units where displacement distances approach or exceed unit dimensions.

We addressed this limitation by aggregating predictions to LGA administrative scales—geographic units sufficiently large (median area approximately 1,800 km²) that most cluster displacements remain within correct jurisdictional boundaries—and by avoiding facility-level or village-scale predictions where positional uncertainty would severely compromise spatial fidelity. Nevertheless, some degree of spatial misclassification persists, particularly in border regions and small LGAs, potentially attenuating observed spatial patterns and introducing noise into administrative unit-level risk estimates.

### 6.3 Diagnostic and Sampling Limitations

Rapid diagnostic test (RDT) technology, while representing substantial advancement over clinical diagnosis, exhibits imperfect sensitivity (85-95%) and specificity (90-98%) that vary by parasite density, *Plasmodium* species composition, and specific RDT product deployed. Diagnostic misclassification introduces measurement error in the outcome variable, with false negatives potentially underestimating risk in low-transmission settings and false positives inflating apparent prevalence. Additionally, the MIS sampling framework—designed to provide nationally and state-representative household samples—systematically underrepresents specific populations at elevated malaria risk including: mobile and migrant populations (agricultural workers, pastoralists, internally displaced persons), institutionalized populations (boarding schools, military barracks, prisons), and geographically remote communities with low population density that may be excluded from sampling frames for logistical or security reasons. These sampling gaps imply that model predictions reflect malaria risk among settled household populations but may not generalize to excluded subpopulations or capture transmission dynamics in conflict-affected or difficult-to-access areas.

### 6.4 Climate and Environmental Variable Limitations

Although the study leverages three decades of gridded climate observations, several constraints limit the environmental component of the predictive framework. First, climate variables were temporally aggregated to align with discrete MIS survey years rather than constructing continuous monthly or seasonal time series, a decision that prioritizes direct linkage between climate exposures and observed outcomes but sacrifices information on sub-annual seasonal dynamics. The timing of survey implementation relative to seasonal transmission peaks—MIS surveys typically occur during a narrow window that may not coincide with peak transmission months—introduces potential temporal misalignment between measured environmental exposures and malaria outcomes. Second, the environmental feature set, while comprehensive, omits potentially relevant covariates including: proximity to water bodies (natural lakes, rivers, irrigation infrastructure) that serve as mosquito breeding sites; land use classifications (agricultural vs. urban vs. forested) that modify microclimate and vector habitat suitability; elevation and topographic indices influencing drainage patterns; and soil moisture data that may better capture breeding site hydrology than rainfall alone. Third, climate data products themselves contain measurement uncertainty, particularly in data-sparse regions where satellite retrievals and reanalysis products have limited ground-station validation.

### 6.5 Absence of Entomological and Health System Indicators

The predictive model focuses exclusively on demographic, environmental, and intervention coverage variables, necessarily omitting entomological and health system factors known to influence malaria transmission and detection. Critical excluded variables include: *Anopheles* species composition and bionomic characteristics (biting behavior, insecticide resistance mechanisms, breeding site preferences) that exhibit substantial geographic heterogeneity across Nigeria; insecticide resistance intensity and mechanisms (metabolic resistance, target-site mutations) that reduce intervention effectiveness; health facility diagnostic testing rates and case management quality that influence detection probability independent of true infection prevalence; healthcare access barriers (travel time to facilities, financial costs, service availability) that determine whether infections are diagnosed; and community-level treatment-seeking behaviors that modify the relationship between infection prevalence and facility-based case reporting. Incorporation of these variables—if systematically collected surveillance data become available—would likely improve model discrimination and provide additional policy-relevant insights regarding intervention optimization.

### 6.6 Outcome Class Imbalance

The distribution of observations across the eight-level malaria risk classification scheme exhibited substantial imbalance, with the majority of clusters concentrated in moderate-risk categories (classes 2-4) and sparse representation of extreme transmission intensity strata (classes 5-7). This class imbalance—reflecting the genuine epidemiological distribution where very high transmission areas are geographically limited—poses challenges for machine learning algorithms that may optimize overall accuracy at the expense of minority class performance. Empirically, the trained Random Forest classifier demonstrated reduced sensitivity (recall) for high-intensity risk classes, meaning that genuinely high-burden areas were sometimes misclassified into lower-risk categories. While stratified sampling and class-weighted loss functions partially mitigate imbalance effects, fundamental data sparsity in extreme transmission zones limits the model’s capacity to learn distinctive patterns characterizing these epidemiologically critical areas. Enhanced surveillance in known high-burden regions or synthetic oversampling techniques might improve minority class performance in future model iterations.

### 6.7 Temporal Non-Stationarity and Model Degradation

Temporal cross-validation revealed pronounced heterogeneity in model performance across survey years, with classification accuracy declining substantially when models trained on earlier surveys were applied to later time periods (2010: 62.2%; 2015: 36.8%; 2021: 40.2%). This temporal performance degradation suggests non-stationarity in the determinants of malaria risk—the statistical relationships between predictor variables and outcomes evolved over the study period in ways not captured by the model’s functional form. Several mechanisms likely contribute to this temporal instability. First, nationwide mass LLIN distribution campaigns implemented between 2009-2015 fundamentally altered the intervention landscape, potentially modifying the predictive importance of climate variables as vector control effectiveness increased. Second, climate variability intensified after 2015, with increased frequency of extreme rainfall events and pronounced droughts that may represent novel climatic conditions outside the environmental parameter space captured in earlier training data. Third, security deterioration and conflict-related population displacement in northeastern states beginning circa 2014 disrupted both malaria control programs and household settlement patterns, introducing unobserved confounding not represented in the predictor set. Fourth, evolution of insecticide resistance mechanisms in vector populations may have reduced the protective effect of ITNs captured in earlier surveys, altering the functional relationship between net ownership and malaria risk.

These sources of temporal non-stationarity underscore the necessity of continual model updating and recalibration as new surveillance data become available, rather than relying on static models trained on historical data. Operational deployment of the predictive framework should incorporate scheduled retraining protocols that incorporate recent epidemiological observations and contemporary climate patterns to maintain predictive accuracy.

### 6.8 Static Behavioral Variables

Intervention coverage variables, particularly ITN usage, were measured through cross-sectional self-reported household survey responses that capture a single point-in-time assessment. This measurement approach cannot capture: temporal variation in net usage across seasons (households may use nets more consistently during peak mosquito seasons); degradation of net physical integrity and insecticidal efficacy over time since distribution; household behavioral dynamics where net ownership does not guarantee consistent usage by all household members; or real-time behavioral responses to perceived malaria risk that may create dynamic feedback relationships between transmission intensity and protective behaviors. More sophisticated measurement approaches incorporating longitudinal behavioral data or sensor-based net usage monitoring could provide time-varying intervention coverage estimates that better capture the dynamic relationship between vector control and transmission.

### 6.9 Limitations for Causal Inference

Fundamentally, the machine learning modeling framework employed prioritizes predictive accuracy and pattern recognition over causal identification. The Random Forest algorithm identifies statistical associations between predictor variables and malaria outcomes but cannot distinguish genuine causal relationships from spurious correlations arising through confounding, reverse causation, or statistical artifacts. Observational survey data—even when enriched with comprehensive environmental covariates—cannot support strong causal claims regarding intervention effectiveness or climate-malaria mechanisms without additional assumptions or study designs (randomized trials, quasi-experimental methods, mechanistic transmission models) that explicitly address confounding and selection bias.

Consequently, policy recommendations derived from model outputs should be viewed as hypothesis-generating rather than definitive evidence of intervention impact, and should be validated through programmatic evaluation studies and targeted field investigations that employ designs better suited to causal inference.

## 7. Conclusion

This study establishes a methodologically transparent and computationally reproducible analytical pipeline integrating nationally representative biomarker surveillance data from Nigeria’s Malaria Indicator Surveys with a comprehensive 34-year hydroclimatic and vegetation time series (1990–2024) to generate spatially explicit malaria risk predictions at policy-relevant administrative scales. The framework addresses critical methodological gaps in existing geospatial malaria modeling approaches by:

1. systematically linking individual-level rapid diagnostic test outcomes with spatiotemporally matched environmental exposures through automated geospatial processing workflows.
2. incorporating climate memory through lagged exposure variables and anomaly metrics that capture biologically plausible delay structures between environmental conditions and malaria transmission.
3. implementing rigorous temporal validation procedures that evaluate predictive performance under operationally realistic scenarios where models must forecast future disease patterns using historical training data.

### 7.1 Principal Findings

The integrated modeling framework yields three significant findings that have direct consequences for malaria control policy and program design. First, the SHAP-based model interpretability analysis found that insecticide-treated net (ITN) coverage was the most important modifiable factor in determining malaria risk. It was much more important than any other environmental variable. This finding provides quantitative validation of vector control as the cornerstone intervention in malaria elimination strategies and suggests that further investments in achieving universal ITN coverage—particularly addressing persistent coverage gaps in northern high-burden states—represent the highest-yield strategy for transmission reduction. The strong predictive signal of ITN coverage also implies that spatially explicit data on intervention deployment can substantially improve risk prediction accuracy, highlighting the value of maintaining high-quality coverage monitoring systems.

Second, climate memory variables—including multi-year lagged rainfall and vegetation indices, long-term climatological baselines, and contemporary anomalies quantifying deviation from expected conditions—contributed meaningfully to predictive performance beyond contemporary climate measurements alone. This result demonstrates that malaria risk reflects not only immediate environmental conditions but also cumulative climate history that shapes vector population dynamics, parasite reservoir prevalence, and human immunity patterns. The predictive value of climate anomalies further suggests opportunities for early warning applications, wherein forecast-based anticipatory actions could be triggered when climate model predictions indicate impending conditions associated with elevated transmission risk.

Third, while the Random Forest ensemble achieved reasonable discrimination capacity (classification accuracy 62.2% for 2010 baseline), temporal cross-validation revealed substantial performance degradation when models trained on earlier surveys were applied to later time periods (36.8% for 2015; 40.2% for 2021). This temporal non-stationarity underscores that malaria risk determinants exhibit evolving relationships over decadal timescales, likely reflecting temporal shifts in intervention coverage following mass distribution campaigns, increasing climate variability with more frequent extreme events, conflict-related population displacement disrupting control programs, and evolution of insecticide resistance reducing intervention effectiveness. These findings emphasize that operational deployment of predictive models requires continuous validation and periodic recalibration as new surveillance data become available, rather than relying on static models with fixed parameters.

### 7.2 Implications for Research and Policy

The demonstrated capacity to integrate multi-decadal climate observations with high-quality biomarker surveillance data offers a scalable template for enhancing spatial targeting of malaria control resources across sub-Saharan Africa. However, translation of predictive models into operational decision support systems requires careful attention to several implementation considerations. First, geographic displacement of household coordinates implemented for privacy protection introduces spatial uncertainty that must be explicitly addressed through aggregation to administrative scales sufficiently large to minimize boundary misclassification. Second, temporal sparsity of biomarker surveys—typically conducted at multi-year intervals—limits the temporal granularity at which models can be validated and constrains confident extrapolation beyond observed time periods. Third, evolving intervention landscapes and non-stationary climate-malaria relationships necessitate institutional mechanisms for regular model updating, requiring sustained data infrastructure, computational capacity, and technical expertise within national malaria control programs.

For policymakers and program implementers, the key operational insight is that sophisticated predictive analytics can meaningfully inform resource allocation decisions when three conditions are met:

1. predictions are generated at administratively actionable spatial scales (Local Government Areas rather than national or pixel-level estimates)
2. model outputs are interpretable and aligned with programmatic levers available to decision-makers (ITN coverage, climate-triggered alerts)
3. predictive systems incorporate feedback loops enabling validation of forecasts against subsequent surveillance data and systematic refinement of model parameters.

When these conditions are satisfied, integration of climate information with epidemiological surveillance transforms malaria control from reactive response to anticipatory programming, enabling preemptive resource prepositioning, optimized geographic targeting, and climate-resilient health system strengthening.

## Data Availability

The data used in this study are publicly available from established repositories. Malaria Indicator Survey (MIS) and Demographic and Health Survey (DHS) datasets for Nigeria were obtained from the DHS Program (https://dhsprogram.com
) upon approved request and are accessible subject to the DHS data use agreement. Climate and environmental datasets, including CHIRPS rainfall data, Normalized Difference Vegetation Index (NDVI), and temperature products, were obtained from open-access sources, including the Climate Hazards Center and Google Earth Engine. All datasets analyzed in this study are anonymized and do not contain personally identifiable information. Derived datasets and machine learning feature outputs generated during the current study are available from the corresponding author upon reasonable request.

https://developers.google.com/earth-engine/datasets

https://developers.google.com/earth-engine/datasets/catalog/MODIS_006_MOD13A2

https://developers.google.com/earth-engine/datasets/catalog/ECMWF_ERA5_LAND_MONTHLY

